# Adults with Adverse Childhood Experiences Report Greater Coronavirus Anxiety

**DOI:** 10.1101/2025.04.09.25325533

**Authors:** Vrinda Kalia, Katherine Knauft

## Abstract

**Background:** Adults with early life adversity exhibit heightened response to threat signals in the environment, which makes them vulnerable to developing stress-related mental health problems, including anxiety disorders. Yet, the impact of the COVID-19 pandemic on this population is understudied. Recently, researchers have characterized dysfunctional cognitions about the pandemic, which are associated with negative mental health outcomes, as coronavirus anxiety.

We conducted a study to examine the relation between exposure to early life adversity, perceived threat from COVID-19, and coronavirus anxiety.

**Methods:** Adults (N = 975; 18-78 years of age; 585 = Women) living in the United States were recruited online in October 2020. Two forms of early life adversity, maltreatment and household dysfunction, were assessed using the Adverse Childhood Experiences scale. Participants’ state anxiety and coronavirus anxiety were measured along with perceived threat from COVID-19. Additionally, as reduced flexibility is implicated in the development and maintenance of anxiety disorders, participants’ cognitive flexibility was assessed using the Cognitive Flexibility Inventory.

**Results:** The data were analyzed using parallel mediation regression analyses. Exposure to early life adversity, in the form of maltreatment and household dysfunction, were the key predictor variables. Coronavirus anxiety and state anxiety were the outcome variables. Perceived threat from COVID-19 and cognitive flexibility were added as parallel mediators into all the regression models. The regression analyses revealed that both perceived threat from COVID-19 and cognitive flexibility mediated the relation between early life adversity and anxiety. The data demonstrate that exposure to early life adversity, in the form of maltreatment or household dysfunction, was associated with higher levels of perceived threat from COVID-19, which, in turn, predicted increased coronavirus anxiety and state anxiety. In contrast, appraisal of everyday challenges as controllable, one of the two types of cognitive flexibility assessed, predicted lower levels of coronavirus anxiety and state anxiety. However, exposure to maltreatment and household dysfunction was associated with reduced cognitive flexibility.

**Conclusion:** This study replicates and extends prior research showing that adults with early life adversity experienced increased anxiety during the pandemic. The findings bolster existing theories that highlight the importance of threat appraisal as a mechanism for the development of anxiety disorders in this population. Additionally, this report adds to the limited body of work on the impact of COVID-19 in adults who have experienced early life adversity.

## Introduction

The COVID-19 pandemic has been unprecedented, with 3.78 million deaths worldwide (600,000 deaths in the US) to date, and has profoundly impacted the mental health of Americans [1]. In the course of an adult’s life, experiences of trauma and stress are to be expected; not everyone encountering these challenges develops enduring mental health problems as a result of these stressors [2]. However, converging streams of data are beginning to show that the pandemic’s impact has been disproportionately negative on vulnerable populations and communities [3]. In particular, individuals in high-risk groups were more likely to experience increased anxiety due to the virus during the pandemic [4].

Adults with early life adversity have been previously identified as vulnerable to stressors [5] yet have received very little attention during the COVID-19 pandemic [6; 7]. Exposure to early life adversity (hereafter; ELA) is implicated in developing a number of health problems later in life [8]. These include increased risk of mental illness, such as anxiety [9], as well as chronic health diseases [10]. Unfortunately, adults who have experienced ELA are not rare in the American population [11]. Per the Centers for Disease Control, approximately 61% (or majority of the adult population) of the surveyed adults in the United States report experiencing at least 1 form of ELA.

### Early life adversity and brain development

Adults who have been exposed to ELA, in the form of abuse, neglect, and household dysfunction prior to 18 years of age, have been identified as vulnerable to stressful experiences in adulthood [5; 12]. One proposed mechanism underlying this vulnerability is the dysregulation of the body’s adaptive acute stress response through extreme and extended activation [5]. The extended activation of the stress response in early development results in the generation of toxic stress, which disrupts the body’s ability to maintain stability in the face of changing environmental demands (or allostasis). Consequently, structural and functional abnormalities in normative brain development, particularly the amygdala and prefrontal cortex (pFC) occur [5].

Empirical evidence demonstrates that ELA alters normative development of the amygdala [13]. Because the amygdala is essential for threat detection, it is more often activated in a home environment that is threatening to the child. Consequently, over time, the amygdala is engaged even in environments that are not stressful [14; 15]. For instance, fMRI research has shown that adults who have been exposed to ELA exhibit heightened amygdala responsiveness when viewing emotional faces in comparison to adults without ELA [16]. Generally, the ability to attend to threats in the environment while attending to ongoing task demands is adaptive [17], as it protects the individual while they carry out goal-directed activities. However, exposure to maltreatment biases attentional and emotional processes toward threatening stimuli in the environment, which results in a sensitized threat system that is specialized to seek out threatening stimuli at the cost of the task at hand [18].

In addition to hyperactive amygdala, adults with ELA have an impaired pFC [5]. For instance, individuals exposed to ELA have smaller frontal brain regions than individuals without ELA [19]. The pFC experiences a protracted developmental period in humans and is flush with glucocorticoid receptors which make it sensitive to stress [5]; consequently, adults with ELA exhibit deficits in cognitive processes that are implicated in activity in the pFC [9]. For example, exposure to ELA in adults is associated with reduced cognitive flexibility [20], which is an executive process that allows an individual to make adaptive behavioral changes [21]. Normally, the amygdala and the pFC would create a network that would allow the individual to evaluate environmental threats effectively [12]. However, adults with ELA have an impaired pFC [5] and a hyperactive amygdala [12], which could enhance their propensity to be emotionally reactive to environmental threats [9].

### Early life adversity as experiences of threat and deprivation

Understanding the impact of early adversity on health outcomes is complicated by the fact that different types of adversities (e.g., abuse, household dysfunction) tend to co-occur [22]. For example, if a child grows up in an abusive home environment they are also more likely to experience neglect from their primary caregivers. As a result, the literature has emphasized studying the cumulative effect of adverse experiences as well as different types of adversity that an individual has experienced [20]. More recently, McLaughlin [23] proposed distinguishing between experiences of deprivation (i.e. the absence of expected environmental inputs that facilitate development) and threat (i.e. the presence of experiences that represent danger) to understand the distinct ways in which different types of ELA could cumulatively impact outcomes.

Using McLaughlin’s model of deprivation and threat [23], Kalia and colleagues [7] studied exposure to ELA in a sample of adults (N = 356). They observed that those adults who had experienced ELA in the form of maltreatment (i.e., threat) perceived a greater threat from COVID-19 in late March 2020, when the US had 85,000 cases of COVID-19 nationwide.

Further, these adults also reported greater state anxiety, which was fully mediated by their perception of the threat posed by COVID-19. These effects remained significant after controlling for the effects of education, SES, age, and gender. No such relationship was observed between perceived threat from COVID-19 and state anxiety in adults who had experienced household dysfunction (i.e., deprivation) early in development. Thus, Kalia and colleagues [7] were able to show that exposure to threatening environments in early development increased reactivity under conditions of threat in adulthood.

## Current Study

We conducted a study to replicate the report by Kalia and colleagues [7]. Additionally, as researchers have recently identified a coherent set of unpleasant cognitions attributed to the fear of COVID-19, named coronavirus anxiety [24], we also sought to extend the work of Kalia and colleagues by examining the relation between ELA and coronavirus anxiety. Coronavirus anxiety is associated with a wide range of psychological issues including elevated depression, generalized anxiety, hopelessness and functional impairments. As such, coronavirus anxiety may be an underlying risk factor for development of mental health problems during the pandemic [25] and after the pandemic is over [26].

Based on the report by Kalia and colleagues [7], our first hypothesis was that exposure to ELA (in the form of maltreatment and household dysfunction) would predict higher state and coronavirus anxiety. Additionally, the relationship between ELA and anxiety (both state anxiety and coronavirus anxiety) would be mediated by perceived threat from COVID-19. Finally, as deficits in cognitive flexibility have been observed in anxiety [27] and early life adversity [20] we also examined the role of cognitive flexibility in the relation ELA and anxiety. Thus, our final hypothesis was that cognitive flexibility would be a parallel mediator between exposure to ELA (in the form of maltreatment and household dysfunction) and anxiety (both state anxiety and coronavirus anxiety).

## Methods

### Participants and procedure

All study procedures were approved by the Institutional Review Board #01620r. In 2020, between October 12^th^ and 14^th^, people (18-78 years; *M_Age_* = 36.28, SD = 10.31, 96.8% were younger than 61 years) living in the United States were recruited via the online platform MTurk to complete the study in exchange for $2.00. Participants provided written informed consent.

After reading the consent form detailing the study participants clicked on either ‘agree’ to participate or ‘disagree’ to participate in the study. Consenting participants’ responses to the consent process were recorded into the survey. At the time of data collection, the United States had more than 7.7 million people who had tested positive for COVID-19 with 40,000-50,000 new cases being reported per day. More than 200,000 Americans had lost their lives to the disease. The sample consisted of 975 participants after excluding 96 participants who failed more than half of the attention checks (Men = 362, Women = 585, Non-binary = 1, Gender Missing = 27).

Majority of the participants identified as white (61.8%), Black or African American (19.1%), Native American (6.5%), Hispanic or Latino (6.3%), Asian or Asian American (3.4%) or preferred not to disclose (3%). The vast majority of participants (95%) had at least some post-high school education, and approximately 25% of the sample held a post-undergraduate degree. To evaluate subjective socioeconomic status, we asked participants to rank themselves from 1 (worst off with the least money, worst employment status) – 10 (best off with the most money, best employment status) on a ladder. Majority (62.3%) of the participants placed themselves at rungs 8 (19.1%), 7 (16.5%), 9 (13.4%), or 6 (13.3%). To evaluate overall health status we used 1 item, *In general would you say your health is* (1 = excellent to 5 = poor), from the 12-item short form health survey [28]. After completing informed consent, participants completed a series of questionnaires that assessed their early life experiences, perceived threat from COVID-19, cognitive flexibility, coronavirus-related anxiety, and state anxiety before completing a demographics form.

### Measures

#### Adverse Childhood Experiences Scale [ACEs; 22]

The ACE scale measures an individual’s exposure to different categories of early life adversity experienced prior to the age of 18. Prior research by [29] has shown the ACE scale measures two underlying factors: maltreatment and household dysfunction. To be consistent with Kalia and colleagues [7], we split the 10-item ACE scale into the maltreatment and a household dysfunction subscale.

Five items of the ACEs scale assess maltreatment (i.e., emotional, physical, and sexual abuse; physical and emotional neglect; *“Did a parent or other adult in the household often swear at you, insult you, put you down, or humiliate you OR act in a way that made you afraid you might be physically hurt”*) and five assess household dysfunction (i.e., domestic violence, parental separation or divorce, substance abuse or mental illness in the home, and incarcerated family members; *“Was a household member depressed or mentally ill or did a household member attempt suicide?”*). Participants respond to each item in a binary fashion (i.e., yes or no). Each yes response is coded as a “1” and “no” responses are coded as “0”. Scores for the two subscales were summed separately. Higher scores on each subscale represent greater exposure to maltreatment (α = .84) or household dysfunction in childhood (α = .85).

#### Perceived Threat of COVID-19

Three items drawn from prior research on pandemics [30] and used by Kalia and colleagues [7] assessed perceived threat. The questions were: 1) How much have you been impacted by COVID-19? (1 = a little to 5 = my life has completely changed); 2) How serious of a problem do you think COVID-19 is? (1 = it’s not very serious to 5 = it is catastrophic); and 3) How likely is it that you would test positive for COVID-19? (1 = not very likely to 5 = extremely likely). Ratings of the three items were summed to create an overall measure of perceived threat from COVID-19.

#### Cognitive Flexibility Inventory [CFI; 31]

We used the CFI to measure self-reported cognitive flexibility. The CFI measures two domains of cognitive flexibility: alternatives and control. The alternatives subscale captures the ability to come up with multiple solutions to a problem or see a problem from multiple perspectives. The control subscale measures the tendency to view challenges as being within one’s control. The CFI contains 20 items that participants respond to via a seven-point Likert scale ranging from *strongly disagree* (1) to *strongly agree* (7). Alternatives subscale is measured through items such as *“I often look at difficult situations from different view-points”* (α = .89) and Control is measured through items such as *“I am capable of overcoming the difficulties in life that I face”* (α = .90). We reverse scored items when necessary and summed responses to create a score for each subscale. Higher scores indicate higher levels of that facet of cognitive flexibility.

#### State-Trait Anxiety Inventory [STAI; 32]

To reduce participant burden, state anxiety was assessed using a 6-item version of the STAI [33]. This version of the STAI has been shown to correlate strongly with the full-length inventory (*r* = .95). Participants respond to items such as *“I feel tense”* on a 4-point Likert scale ranging (1 = *not at all* to 4 = *very much so*). Scores are summed to create an overall score, with higher scores indicating greater state anxiety (α = .74).

#### Coronavirus Anxiety Scale [CAS; 24]

Coronavirus anxiety was assessed using a five-item scale that was designed to identify COVID-related anxiety that reaches the level of dysfunction or impairment. The items ask participants about negative experiences over the last two weeks related to information or thoughts about coronavirus, such as *“I felt paralyzed or frozen when I thought about or was exposed to information about the coronavirus”*. Each item is rated on a Likert scale (0 = *not at all* to 4 = *nearly every day over the last two weeks*). Scores are summed, with higher scores indicating more anxiety related to COVID-19 (α = .92). Scores greater than 9 are thought to indicate levels of coronavirus anxiety that are dysfunctional [24]. Within our sample, 46.2% met or exceeded this threshold, suggesting that these individuals experienced levels of coronavirus anxiety that may be dysfunctional.

## Data Analytic Plan

Parallel mediation regression models will be conducted using Hayes’ PROCESS 3 macro model 4 [34] in SPSS version 25.0. ACEs-maltreatment or ACEs-household dysfunction will serve as the independent variables in the models with state anxiety or coronavirus anxiety as the dependent variables. To be consistent with Kalia and colleagues [7], CFI-Control and perceived threat from COVID-19 will be tested as parallel mediators (See Figure 1). Across all 4 models participant’s age, gender, subjective SES, education, overall health status, and race/ethnicity will be added as covariates.

**Figure 1.**
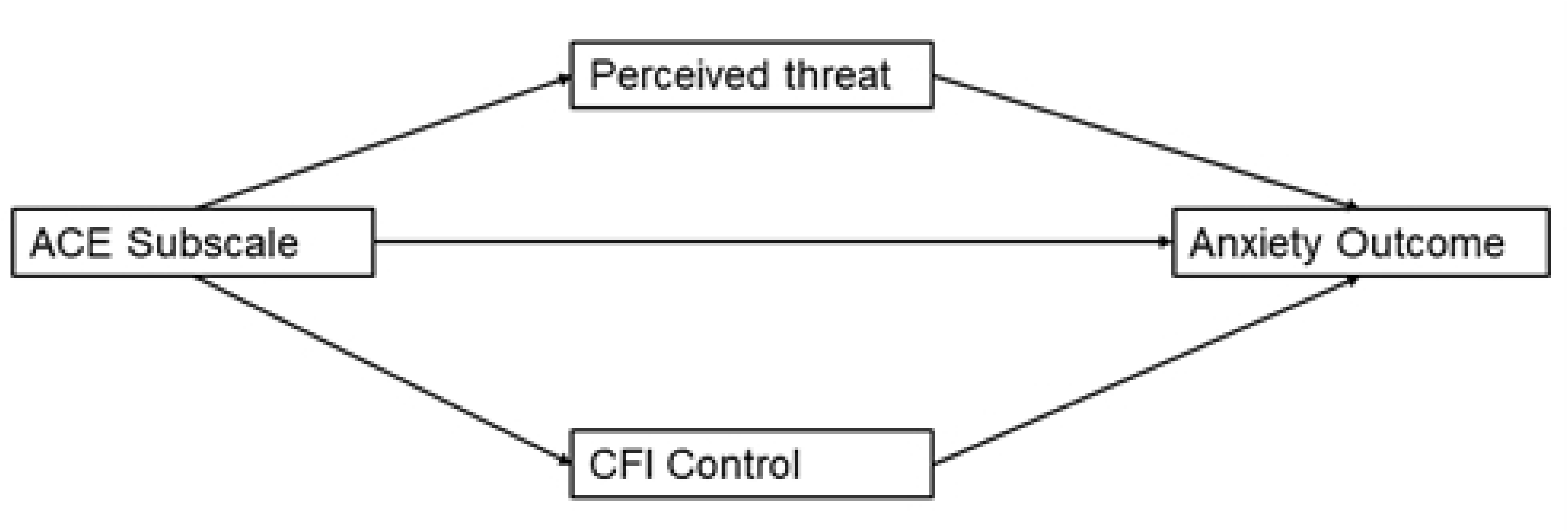
Conceptual model of proposed parallel mediation.

## Results

### Descriptive Statistics and Correlations

Data were normally distributed (skew coefficient < |1|; kurtosis coefficient < |2|).

Bivariate correlations and descriptive statistics are presented in Table 1. Consistent with prior studies [e.g., 20; 7], approximately 63% of participants reported experiencing at least one form of maltreatment and 52% of participants reported experiencing at least one form of household dysfunction (See Table 2 and Figure 2).

**Figure 2.**
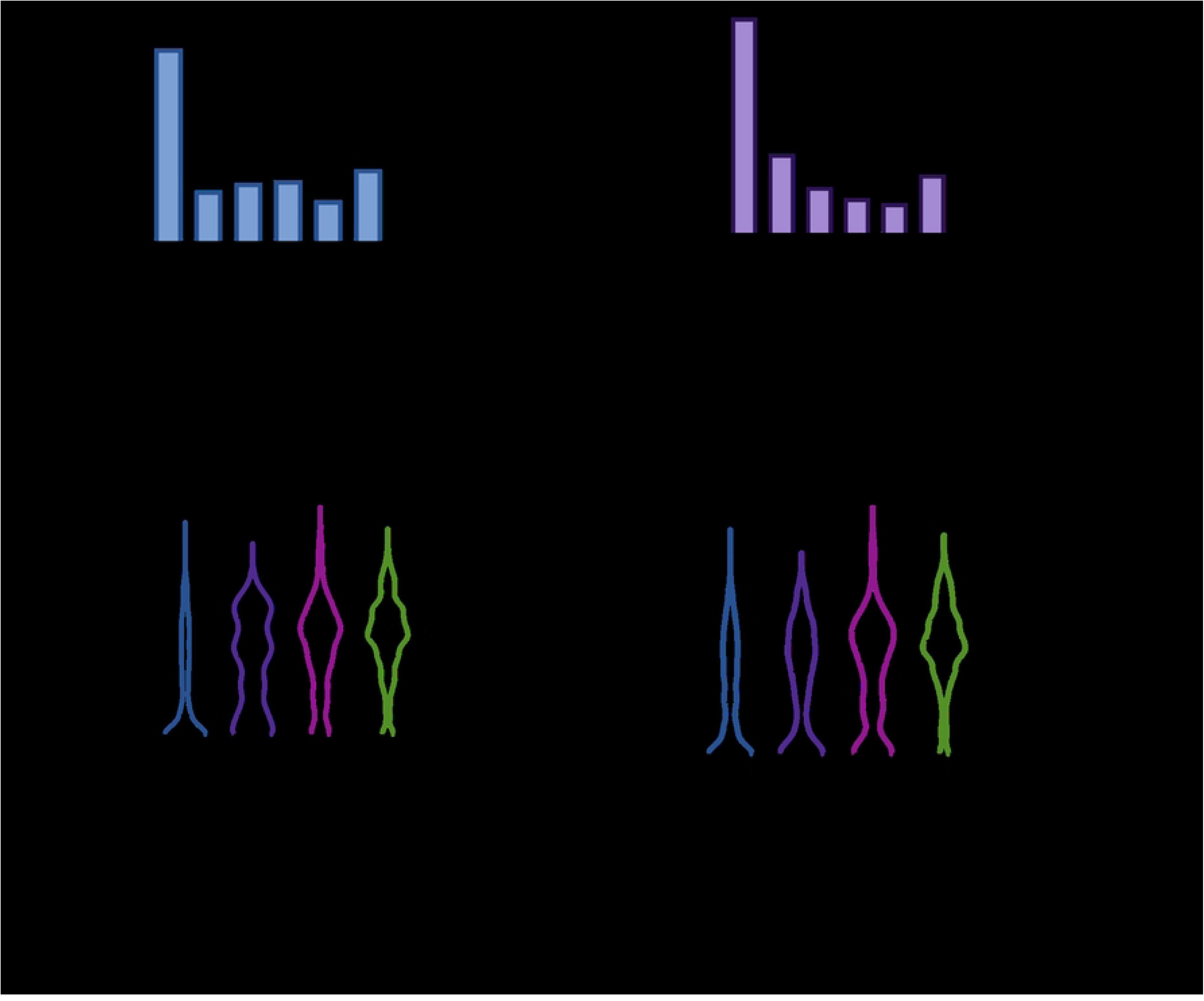
Frequencies of reported ACE-Maltreatment (A) and ACE-Household Dysfunction scores; Violin plots of associations between coronavirus anxiety and ACE-Maltreatment Scores (C) and ACE-Household Dysfunction scores (D); Within violin plots the center of each distribution is the median, which can be identified by the thicker line with larger dashes. The 25^th^ and 75^th^ quartiles can be identified by the thinner lines with smaller dashes.

**Table 1.** Correlations and descriptive statistics.

|  | 1. | 2. | 3. | 4. | 5. | 6. | 7. | 8. | 9. | 10. | 11. |
| --- | --- | --- | --- | --- | --- | --- | --- | --- | --- | --- | --- |
| 1. ACE-Maltreatment | - |  |  |  |  |  |  |  |  |  |  |
| 2. ACE-Household Dysfunction | .81*** | - |  |  |  |  |  |  |  |  |  |
| 3. Perceived Threat | .14*** | .10** | - |  |  |  |  |  |  |  |  |
| 4. CFI Control | -.41*** | -.32*** | -.27*** | - |  |  |  |  |  |  |  |
| 5. CFI Alternatives | .06 | .08* | .18*** | .001 | - |  |  |  |  |  |  |
| 6. Coronavirus Anxiety | .47*** | .39*** | .42*** | -.70*** | .06* | - |  |  |  |  |  |
| 7. State Anxiety | .37*** | .36*** | .21*** | -.56*** | -.17*** | .49*** | - |  |  |  |  |
| 8. Age | -.03 | .01 | -.01 | .12*** | .06 | -.10** | -.06 | - |  |  |  |
| 9. SES | .29*** | .19*** | .15*** | -.45*** | .03 | .51*** | .17*** | -.04 | - |  |  |
| 10. Education | .11** | .02 | .15*** | -.29*** | .04 | .30*** | .12*** | -.08* | .41*** | - |  |
| 11. General Health | -.14*** | -.15*** | .05 | .05 | .18*** | .02 | -.27*** | -.01 | .21*** | .15*** | - |
| Mean | 1.84 | 1.49 | 8.42 | 26.25 | 71.87 | 7.25 | 12.39 | 36.28 | 6.99 | 3.97 | 3.72 |
| SD | 1.88 | 1.80 | 2.29 | 9.22 | 9.39 | 5.31 | 3.54 | 10.31 | 2.05 | 1.01 | 0.86 |

*Note.* \* $p < .05$ ; \*\* $p < .01$ ; \*\*\* $p < .001$ ; Education: 1 = Some high school, 8 = Doctorate (PhD, MD, etc); The final dataset had 944 participants with information on state anxiety and 937 participants with information on coronavirus anxiety.

**Table 2.** Frequencies of adverse childhood experiences.

| Adverse Childhood Experiences | Frequency (%) |  |  |
| --- | --- | --- | --- |
|  | Women (n = 362) | Men (n = 585) | Total (N = 975) |
| Emotional abuse | 152 (42.0%) | 262 (44.8%) | 415 (42.6%) |
| Physical abuse | 116 (32.0%) | 214 (36.6%) | 330 (33.8%) |
| Sexual abuse | 111 (30.7%) | 183 (31.1%) | 293 (30.1%) |
| Emotional neglect | 122 (33.7%) | 223 (38.1%) | 345 (35.4%) |
| Physical neglect | 135 (37.3%) | 236 (40.3%) | 372 (38.2%) |
| Parental separation or divorce | 114 (31.5%) | 176 (30.1%) | 290 (29.7%) |
| Domestic violence against parent | 95 (26.2%) | 189 (32.3%) | 284 (29.1%) |
| Household alcohol/drug abuse | 113 (31.2%) | 209 (35.7%) | 322 (33.0%) |
| Mental illness in household | 99 (27.3%) | 178 (30.4%) | 277 (28.4%) |
| Incarcerated family member | 79 (21.8%) | 164 (28.4%) | 243 (24.9%) |
| **Adverse Childhood Experiences Score** |  |  |  |
| 0 | 119 (32.9%) | 190 (32.5%) | 312 (32.0%) |
| 1 | 50 (13.8%) | 57 (9.7%) | 108 (11.1%) |
| 2 | 28 (7.7%) | 55 (9.4%) | 83 (8.5%) |
| ≥ 3 | 164 (45.3%) | 282 (48.3%) | 446 (45.7%) |

Note. 28 participants were non-binary or did not disclose their gender

### 3.2. Parallel mediation models with maltreatment as the predictor variable

In both models, maltreatment and the covariates accounted for 5% of the variance in perceived threat, *F*(7, 936) = 6.41, *p* < .001. Maltreatment, *B* = 0.14, *t*(936) = 3.40, *p* < .001, and education, *B* = 0.24, *t*(936) = 2.95, *p* = .003, emerged as significant predictors of increased threat perception. Maltreatment and the covariates accounted for 32% of the variance in CFI-Control, *F*(7, 936) = 63.08, *p* < .001. Maltreatment, *B* = -1.40, *t*(936) = -9.83, *p <* .001, age, *B* = 0.08, *t*(936) = 3.15, *p* = .002, education, *B* = -1.26, *t*(936) = -4.60, *p* < .001, SES, *B* = -1.46, *t*(936) = -10.26, *p* < .001, and general health, *B* = 0.98, *t*(936) = 3.29, *p* = .001, emerged as significant predictors of CFI-Control. Increased exposure to maltreatment, higher education and subjective SES predicted lower scores on CFI-Control, whereas older and healthier participants had higher scores on CFI-Control.

#### 3.2.1. Maltreatment predicts state anxiety

Maltreatment, the two mediators, and the covariates accounted for 40% of the variance in state anxiety, *F*(9, 934) = 68.28, *p* < .001. Both perceived threat, *B* = 0.12, *t*(934) = 2.97, *p* = .003, and CFI-Control, *B* = -.18, *t*(934) = -15.41, *p* < .001, emerged as significant mediators. Higher perceived threat was associated with increased state anxiety, whereas higher scores on CFI-Control were associated with lower state anxiety. The total effect of maltreatment on state anxiety (excluding mediators) was significant, *B* = 0.54, *t*(936) = 9.32, *p* < .001. When mediators were included in the model, the direct effect of maltreatment on state anxiety remained significant, *B* = 0.27, *t*(934) = 4.96, *p* < .001. A 95% bias-corrected confidence interval based on 10,000 bootstrapped samples indicated that the indirect effect through threat perception, *B* = .02, *SE* = .03, 95% CI: [.004, .036] was above zero. Likewise, the indirect through CFI-Control, *B* = .26, *SE* = .03, 95% CI: [.20, .32] was above zero. Thus, both indirect effects were significant (See Figure 3).

**Figure 3.**
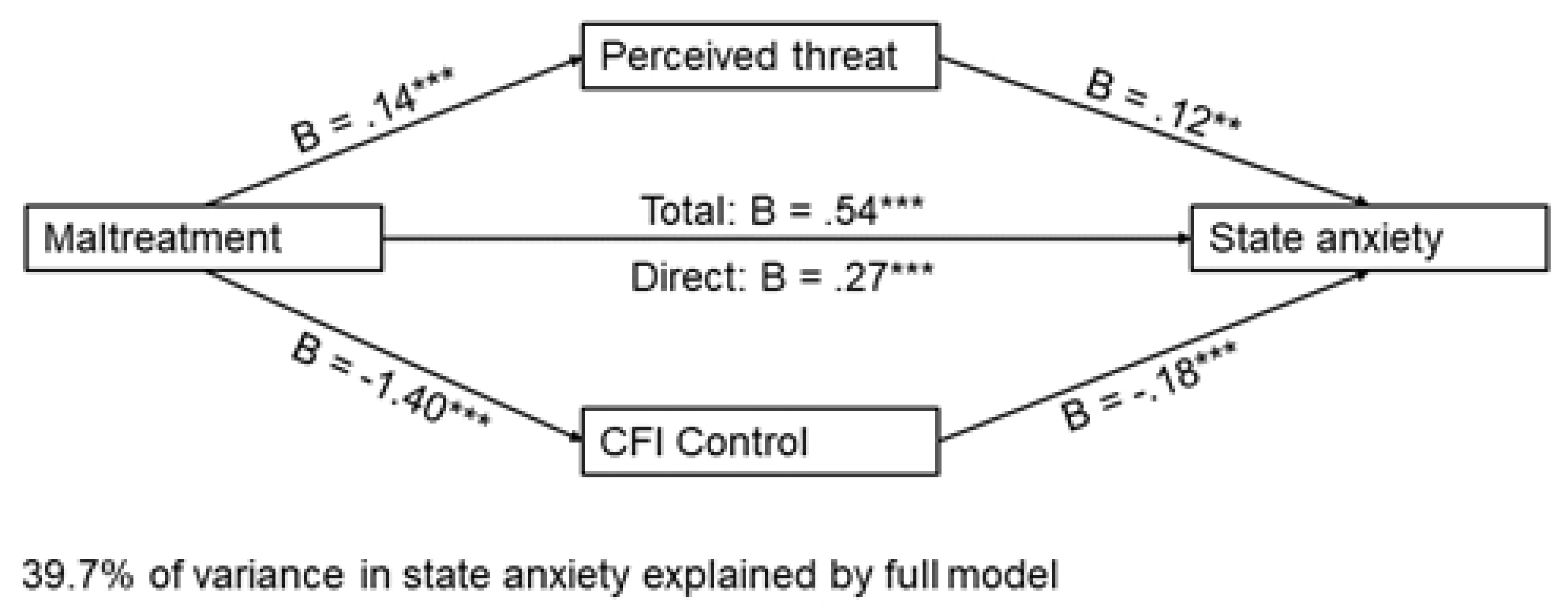
Parallel mediation model with ACE-Maltreatment as the independent variable and state anxiety as the dependent variable. *Note.* Figure represents data from n = 944 participants

#### 3.2.2. Maltreatment predicts coronavirus anxiety

Maltreatment, the mediators, and the covariates accounted for 62% of the variance in coronavirus anxiety, *F*(9, 927) = 171.42, *p* < .001. Both perceived threat, *B* = 0.52, *t*(927) = 10.65, *p* < .001, and CFI-Control, *B* = -.27, *t*(927) = -18.76, *p* < .001, emerged as significant mediators. Higher perceived threat was associated with increased coronavirus anxiety, whereas elevated CFI-Control was associated with lower scores on the coronavirus anxiety scale. The total effect of maltreatment on coronavirus anxiety (excluding mediators) was significant, *B* = 0.99, *t*(929) = 12.74, *p* < .001. When mediators were included in the model, the direct effect of maltreatment on coronavirus anxiety remained significant, *B* = 0.53, *t*(927) = 8.27, *p* < .001. A 95% bias-corrected confidence interval based on 10,000 bootstrapped samples indicated that the indirect effect through threat perception, *B* = .08, *SE* = .02, 95% CI: [.033, .127] was above zero. Likewise, the indirect through CFI-Control, *B* = .38, *SE* = .04, 95% CI: [.30, .466] was above zero. Thus, both indirect effects were significant (See Figure 4).

**Figure 4.**
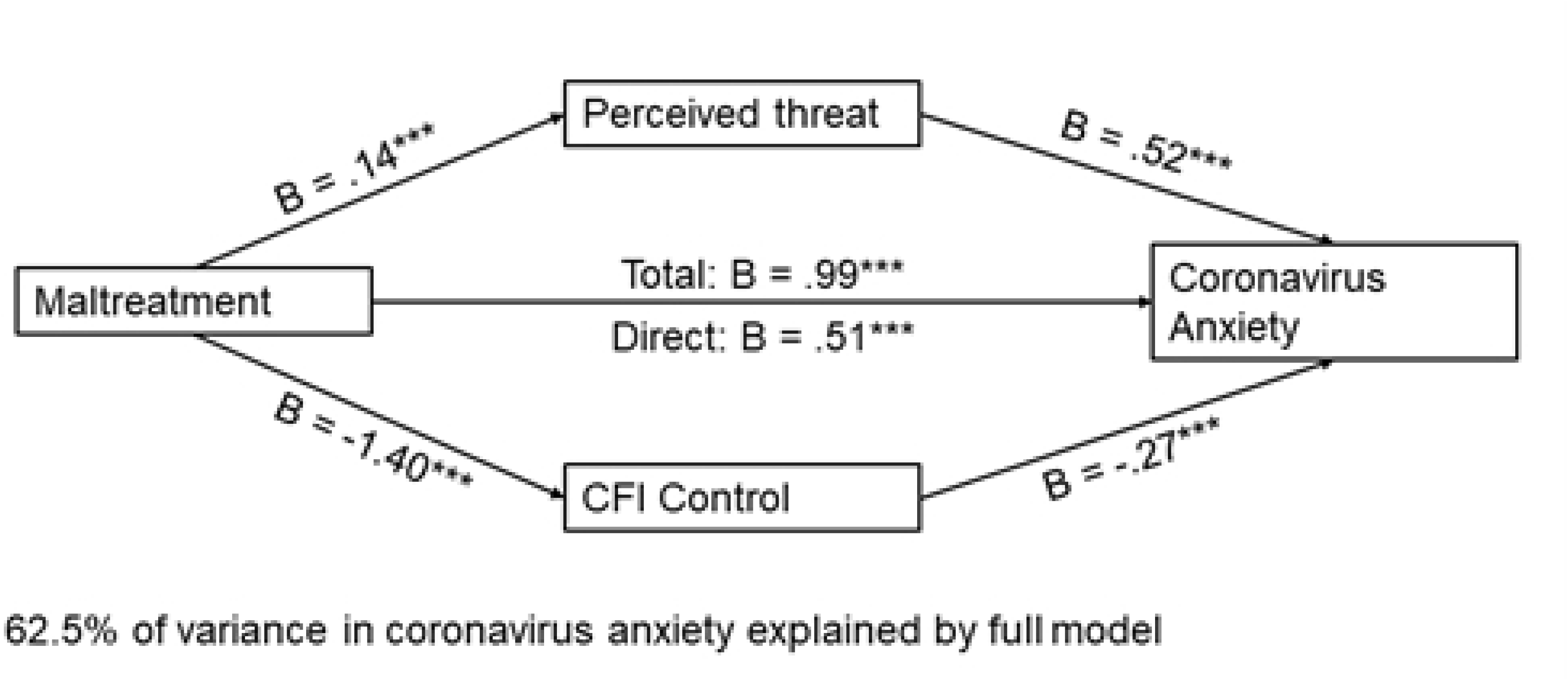
Parallel mediation model with ACE-Maltreatment as the independent variable and coronavirus anxiety as the dependent variable. *Note.* Figure represents data from n = 937 participants

### 3.3. Parallel mediation models with household dysfunction as the predictor variable

In both models, household dysfunction and the covariates accounted for 4% of the variance in perceived threat, *F*(7, 936) = 5.66, *p* < .001. Household dysfunction, *B* = 0.11, *t*(936) = 2.57, *p* = .010, education, *B* = 0.25, *t*(936) = 3.08, *p* = .002, and SES, *B* = 0.09, *t*(936) = 2.31, *p* = .021, emerged as significant predictors of increased threat perception. Household dysfunction and the covariates accounted for 30% of the variance in CFI-Control, *F*(7, 936) = 56.89, *p* < .001. Household dysfunction, *B* = -1.17, *t*(936) = -7.98, *p <* .001, age, *B* = 0.08, *t*(936) = 3.32, *p* < .001, education, *B* = -1.36, *t*(936) = -4.87, *p* < .001, subjective SES, *B* = -1.62, *t*(936) = -11.48, *p* < .001, and general health, *B* = 1.13, *t*(936) = 3.77, *p* < .001, emerged as significant predictors of CFI-Control. More household dysfunction, higher education and subjective SES predicted lower scores on CFI-Control, whereas older and healthier participants had higher scores on CFI-Control.

#### 3.3.1. Household dysfunction predicts state anxiety

Household dysfunction, the mediators, and the covariates accounted for 41% of the variance in state anxiety, *F*(9, 934) = 71.06, *p* < .001. Both perceived threat, *B* = 0.12, *t*(934) = 3.01, *p* = .003, and CFI-Control, *B* = -.18, *t*(934) = -15.72, *p* < .001, emerged as significant mediators. Higher perceived threat was associated with increased state anxiety, whereas higher scores on CFI-Control were associated with lower state anxiety. The total effect of household dysfunction on state anxiety (excluding mediators) was significant, *B* = 0.57, *t*(936) = 9.61, *p* < .001. When mediators were included in the model, the direct effect of maltreatment on state anxiety remained significant, *B* = 0.34, *t*(934) = 6.32, *p* < .001. A 95% bias-corrected confidence interval based on 10,000 bootstrapped samples indicated that the indirect effect through threat perception, *B* = .01, *SE* = .01, 95% CI: [.002, .031] was above zero. Likewise, the indirect through CFI-Control, *B* = .22, *SE* = .03, 95% CI: [.17, .27] was above zero. Thus, both indirect effects were significant (See Figure 5).

**Figure 5.**
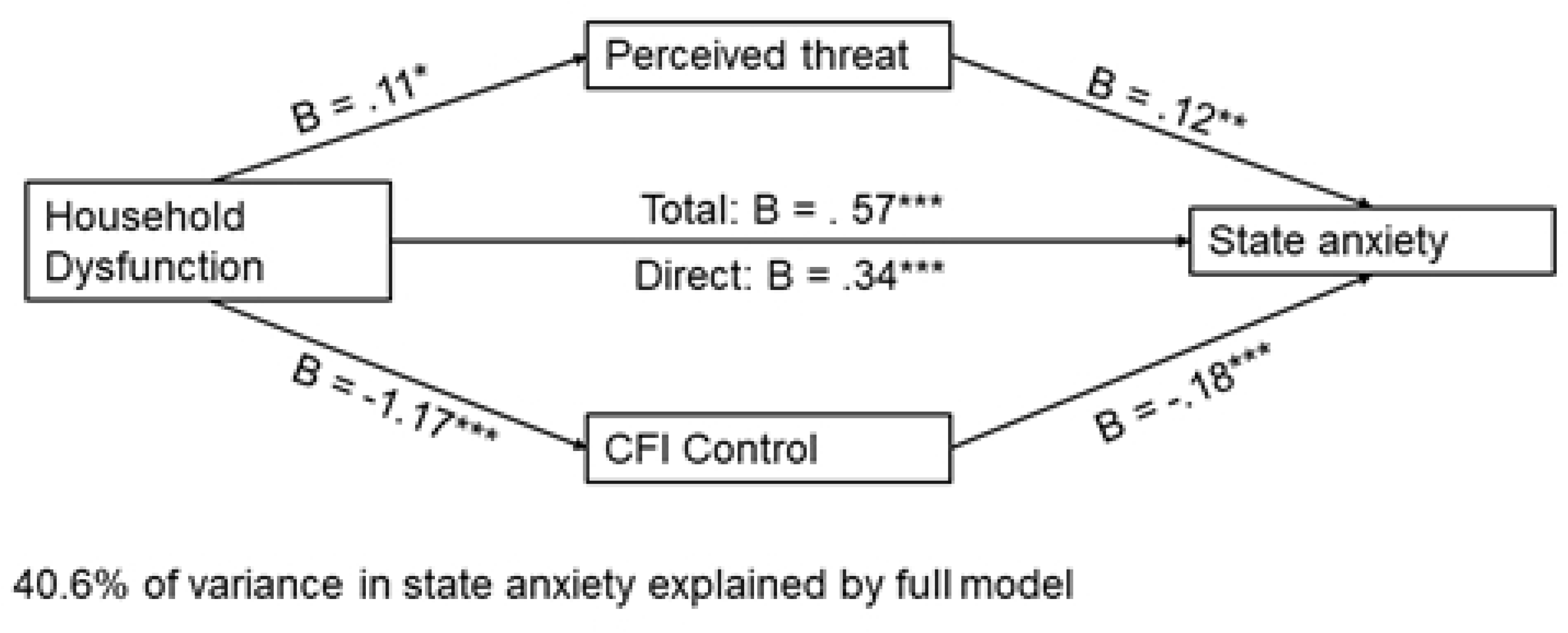
Parallel mediation model with ACE-Household Dysfunction as the independent variable and state anxiety as the dependent variable. *Note.* Figure represents data from n = 944 participants

#### 3.3.2. Household dysfunction predicts coronavirus anxiety

Household dysfunction, the mediators, and the covariates accounted for 62% of the variance in coronavirus anxiety, *F*(9, 927) = 171.08, *p* < .001. Both perceived threat, *B* = 0.53, *t*(927) = 10.82, *p* < .001, and CFI-Control, *B* = -.27, *t*(927) = -19.46, *p* < .001, emerged as significant. Higher perceived threat was associated with more coronavirus anxiety, while elevated CFI-Control was associated with lower coronavirus anxiety. The total effect of household dysfunction on coronavirus anxiety was significant, *B* = 0.91, *t*(929) = 11.41, *p* < .001. When mediators were included in the model, the direct effect of maltreatment on coronavirus anxiety remained significant, *B* = 0.52, *t*(927) = 8.20, *p* < .001. A 95% bias-corrected confidence interval based on 10,000 bootstrapped samples indicated that the indirect effect through threat perception, *B* = .06, *SE* = .02, 95% CI: [.016, .110] was above zero.

Likewise, the indirect through CFI-Control, *B* = .32, *SE* = .04, 95% CI: [.25, .40] was above zero. Thus, both indirect effects were significant (See Figure 6).

**Figure 6.**
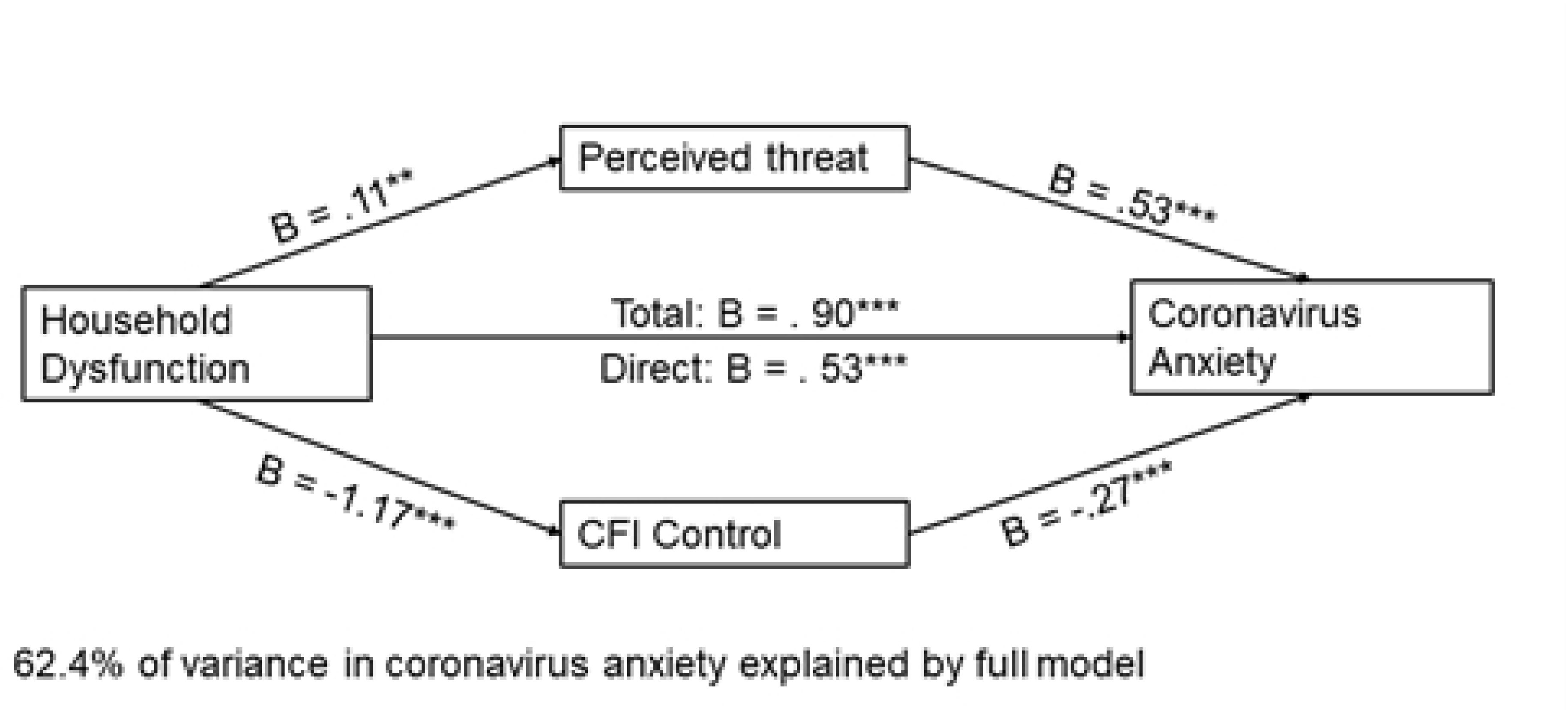
Parallel mediation model with ACE-Household Dysfunction as the independent variable and coronavirus anxiety as the dependent variable. *Note.* Figure represents data from n = 937 participants

### 4. Discussion

The results of the mediation regression analyses demonstrated that maltreatment and household dysfunction predicted increased state anxiety and coronavirus anxiety via greater perceived threat of COVID-19 and reduced cognitive flexibility. Our data replicate the work of Kalia and colleagues [7] who observed that enhanced perceived threat and reduced cognitive flexibility mediated the relationship between ACE-maltreatment and state anxiety. These findings are also consistent with other reports showing that adults with ELA have experienced psychological distress during the COVID-19 global pandemic [35; 36].

Intriguingly, the presented data show that both ACE-maltreatment and ACE-household dysfunction predict increased state anxiety whereas Kalia and colleagues [7] only observed a significant association between ACE-maltreatment and state anxiety. This difference may be due to the timing of data collection, as Kalia and colleagues collected their data in March 2020 when the US population was split in their appraisal of the threat posed by the pandemic [37]. Since adults with exposure to early life maltreatment exhibit enhanced neural response to environmental threats even when they are ambiguous [38], it is plausible that the anxiety that maltreated adults reported in the early part of the pandemic is due to a vulnerability conferred specifically because of the impact of maltreatment on the development of the amygdala-prefrontal circuitry [14; 12].

However, the pandemic had dramatically expanded and intensified by October 2020 when we collected data for this study. Thus, it is possible that the observed association between ELA and anxiety is a consequence of the sustained activation of the stress systems leading to increased allostatic load [5]. One interesting proposal is that long periods of intense stress disrupt the excitation-inhibition balance of the amygdala that was already vulnerable because of exposure to early life stress [39]. Since we did not conduct a neuroimaging study we cannot address this issue directly. However, the fact that enhanced activity in the amygdala is associated with anxiety [40], and perceived threat partially mediated the relation between ELA and anxiety in our work should provide impetus for human neuroscience research to systematically study the impact of the pandemic on the amygdala-prefrontal circuitry in adults with ELA.

As we did not collect data on trait levels of anxiety, we cannot make any claims about an individual’s propensity to experience anxiety [41]; but, we found that both maltreatment and household dysfunction are associated with increased state anxiety and coronavirus anxiety during the pandemic. This may suggest that studying subtypes of adversity is insightful; but perhaps less informative about differential relations with outcomes in the midst of an extremely stressful chronic life event (e.g., global pandemic). Thus, our work provides some support for the notion that both experiences of threat and deprivation present themselves as questions of survival [42], such that in adults who have experienced ELA it may increase the probability of enhanced reactivity during stressful experiences.

We also examined coronavirus anxiety in relation to ELA. The coronavirus anxiety scale (CAS) captures dysfunctional anxiety associated with COVID-19, and high scores on the CAS are associated with functional impairment and distress [24; 25). The relation between ELA and coronavirus anxiety was partially mediated by perceived threat of COVID-19. Adults with ELA were more likely to report greater threat from COVID-19 which partially accounted for their higher scores on the coronavirus anxiety scale. Since the amygdala is implicated in threat processing [43], our data are concordant with neuroimaging research showing that increased sensitivity of the amygdala to threat may mediate anxiety in adults with ELA [40]. To the best of our knowledge, we are the first to observe that adults with ELA also experienced increased levels of coronavirus anxiety. Considering that adults with ELA are vulnerable to developing anxiety disorders [40] and cognitions about COVID-19 may predict persistent negative mental health outcomes past the pandemic [26], our work suggests the need for further research on the relation between ELA and coronavirus anxiety. Despite being adequately powered, this novel finding must be replicated before any firm claims can be made about the relationship between ELA and coronavirus anxiety.

Consistent with Kalia and colleagues [7] we observed that exposure to ELA was associated with reduced cognitive flexibility. Additionally, cognitive flexibility partially mediated the association between ELA and anxiety, such that lower levels of flexibility predicted higher levels of anxiety. Our results provide some support for the notion that deficits in cognitive flexibility are associated with anxiety [27]. Because cognitive flexibility can be enhanced via exposure to enriching environmental experiences [21], future research should examine whether increasing cognitive flexibility can aid with reducing anxiety in adults with ELA.

Since scores on CFI-Control are an indicator of an individual’s ability to appraise everyday challenges as controllable [31], our findings indicate that adults with ELA are more likely to view future challenges as uncontrollable, which increases their anxiety. This is concordant with the proposal that experiencing a lack of control over future events (i.e., everyday challenges) precludes an individual from feeling that they have the resources to avert an aversive event, which can enhance anxiety [44]. Our data were collected in the midst of a global pandemic, so it is relevant to point out that feeling some anxiety during stressful events can be adaptive as it appropriately allows attention to focus on the threat in the environment [44]. However, as seen in Figure 2, adults with 2 or more ACE-maltreatments and 3 or more ACE-household dysfunction had maladaptive levels (i.e., scores higher than 9) [24] of coronavirus anxiety.

Overall, our results suggest that adults with ELA are vulnerable to anxiety during stressful circumstances because they experience a greater degree of threat from an environmental stressor and are less likely to feel in control during the stressful event.

### 4.1. Limitations and Conclusions

There are several limitations that must be acknowledged when interpreting our findings. First, our sample size is large but not representative of the population of the United States (e.g., disproportionately female and younger). Additionally, our data collection was restricted to individuals living in the United States and the findings may not generalize to other countries.

Second, it is important to point out that the ACE scale measures early life adversity retrospectively which makes it susceptible to errors of recall. Prospective longitudinal studies should be conducted to fully understand the relationship between early life adversity and coronavirus anxiety. Third, we used a self-report measure of cognitive flexibility so our results may not extend to behavioral measures of cognitive flexibility. Though all the measures we used are either well-established (e.g., state anxiety) or have been used for replication (i.e. perceived threat), our data are correlational in nature and emerged from self-report measures; thus no causal claims can be made. Nevertheless, our work demonstrates that adults with ELA are vulnerable to debilitating anxiety during the pandemic and should be carefully considered when the psychological toll of COVID-19 is evaluated.

## Data Availability

Due to the sensitive nature of the data, the IRB has not provided permission to share the data openly. Please email the first author for deidentified data for research purposes only.

## Notes

### Competing Interest Statement

The authors have declared no competing interest.

### Funding Statement

The author(s) received no specific funding for this work.

### Author Declarations

Department of Psychology Review Board, Miami University

